# Validation of an AI-based solution for breast cancer risk stratification using routine digital histopathology images

**DOI:** 10.1101/2023.10.10.23296761

**Authors:** Abhinav Sharma, Sandy Kang Lövgren, Kajsa Ledesma Eriksson, Yinxi Wang, Stephanie Robertson, Johan Hartman, Mattias Rantalainen

## Abstract

**Background:** Stratipath Breast is a CE-IVD marked artificial intelligence-based solution for prognostic risk stratification of breast cancer patients into high- and low-risk groups, using haematoxylin and eosin (H&E)-stained histopathology whole slide images (WSIs). In this validation study, we assessed the prognostic performance of Stratipath Breast in two independent breast cancer cohorts.

**Methods:** This retrospective multi-site validation study included 2719 patients with primary breast cancer from two Swedish hospitals. The Stratipath Breast tool was applied to stratify patients based on digitised WSIs of the diagnostic H&E-stained tissue sections from surgically resected tumours. The prognostic performance was evaluated using time-to-event analysis by multivariable Cox Proportional Hazards analysis with progression-free survival (PFS) as the primary endpoint.

**Results:** In the clinically relevant oestrogen receptor (ER)-positive/human epidermal growth factor receptor 2 (HER2)-negative patient subgroup, the estimated hazard ratio (HR) associated with PFS between low- and high-risk groups was 2.76 (95% CI: 1.63-4.66, p-value < 0.001) after adjusting for established risk factors. In the ER+/HER2-Nottingham histological grade (NHG) 2 subgroup, the HR was 2.20 (95% CI: 1.22-3.98, p-value = 0.009) between low- and high-risk groups.

**Conclusion:** The results indicate an independent prognostic value of Stratipath Breast among all breast cancer patients, as well as in the clinically relevant ER+/HER2-subgroup and the NHG2/ER+/HER2-subgroup. Improved risk stratification of intermediate-risk ER+/HER2-breast cancers provides information relevant for treatment decisions of adjuvant chemotherapy and has the potential to reduce both under- and overtreatment. Image-based risk stratification provides the added benefit of short lead times and substantially lower cost compared to molecular diagnostics and therefore has the potential to reach broader patient groups.

## Introduction

In breast cancer, Nottingham histological grade (NHG) is a well-established prognostic factor that provides information for clinical decision-making (1,2). Patients with oestrogen receptor (ER)-positive/human epidermal growth factor receptor 2 (HER2)-negative tumours with high-risk clinical factors such as high histological grade (i.e. NHG3) are often considered for adjuvant chemotherapy whereas patients whose tumors are associated with low-risk clinical factors (i.e. NHG1) can be spared chemotherapy. However, more than 50% of patients belong to the intermediate risk category, NHG2, of limited clinical value (3). Consequently, many patients may be overtreated with considerable side effects or undertreated with risk of recurrence.

Several multigene expression-based methods have been developed to predict the risk of recurrence for ER+ early-stage breast cancer patients (4–6), in particular within the intermediate risk category. The most commonly used products include MammaPrint (Agendia Inc., Amsterdam, The Netherlands) (7), Oncotype DX (Exact Sciences Corp., Madison, WI, USA) (8), EndoPredict/EPclin (Myriad Genetics Inc., Salt Lake City, UT, USA) (9) and Prosigna (Veracyte Inc., South San Francisco, CA, USA) (10). Oncotype DX recurrence score (RS) was one of the first multigene assay-based tests to predict the distant recurrence in ER+ node-negative tamoxifen-treated breast cancer patients by stratifying patients into low-, intermediate- and high-risk groups (8). Prosigna ROR score has been validated to predict the distant and late-distant (5-10 years) recurrence in postmenopausal hormone receptor-positive early breast cancer patients (4,10,11). Furthermore, models based on molecular signatures, like the Genomic grade index (GGI) (12), have been proposed to improve the stratification of intermediate-grade patients (13). However, in clinical practice, the use of molecular multigene assays implies long lead times and remains expensive (14).

Enabled by the emergence of digital and computational pathology, deep learning-based whole slide image (WSI) classification models have recently been demonstrated to enable improved prognostic stratification of patients compared to routine pathology, and to enable faster and more precise information for clinical decision-making in several cancer types (14–18). Stratipath Breast (Stratipath AB, Stockholm, Sweden) is the first CE-IVD marked artificial intelligence (AI)-based image analysis tool for primary breast cancer risk stratification that is available for routine clinical use. Stratipath Breast provides risk stratification of breast cancer patients into low- and high-risk groups based on the assessment of routine haematoxylin and eosin (H&E)-stained tumour tissue slides from surgical resection specimens. In clinical practice, risk stratification of intermediate-risk ER+/HER2-patients is of particular interest, as improved prognostic information can provide guidance relating to treatment decisions for adjuvant chemotherapy, and thus reduce over- and undertreatment of patients. Stratipath Breast only requires an H&E-stained WSI from the resected tumour as input and uses a deep learning-based model to risk-stratify patients into low- and high-risk groups. It is of clinical importance that the prognostic performance and generalisability of AI-based tools for decision support are validated in independent study materials to ensure that the solution provides significant prognostic value and that it generalises well across sites. In this study, we validated the prognostic performance of Stratipath Breast in two fully independent breast cancer cohorts from two sites in Sweden.

## Methods

### Study Materials

In this study, we included breast cancer patients from two Swedish studies: the CHIME breast KS Solna cohort and the SCAN-B (Lund) study. CHIME breast KS Solna is a population representative retrospective cohort including primary breast cancer patients (N=2922) from the Karolinska University Hospital Solna, Stockholm, diagnosed between 2009-2018. Clinicopathological information was retrieved from the National Quality Registry for Breast Cancer (NKBC). The SCAN-B (Lund) study includes a subset of the patients (N=1262) from the prospective SCAN-B study (19), diagnosed between 2010-2019 in Lund, Sweden. Archived formalin-fixed paraffin-embedded H&E-stained tissue slides were retrieved and scanned at 40x magnification using Hamamatsu NanoZoomer XR or S360 whole slide scanner (Hamamatsu Photonics K.K., Shizuoka, Japan) and Aperio GT 450 DX Digital Pathology slide scanner (Leica Biosystems, Wetzlar, Germany). One H&E-stained tumour WSI for each patient was used for analyses. Patients with missing clinical information were excluded from the analyses. Patients diagnosed with invasive breast cancer without receiving neoadjuvant treatment were included in this study and only WSIs from resection specimens were included. In total, 102 WSIs were used for the Stratipath Breast system setup procedure, and these were excluded from further analyses. All exclusion criteria are described in the CONSORT diagram in Figure 1. A total of 2719 patients were included for further analyses and the baseline characteristics of the cohorts are available in Supplementary Table 1.

**Figure 1.**
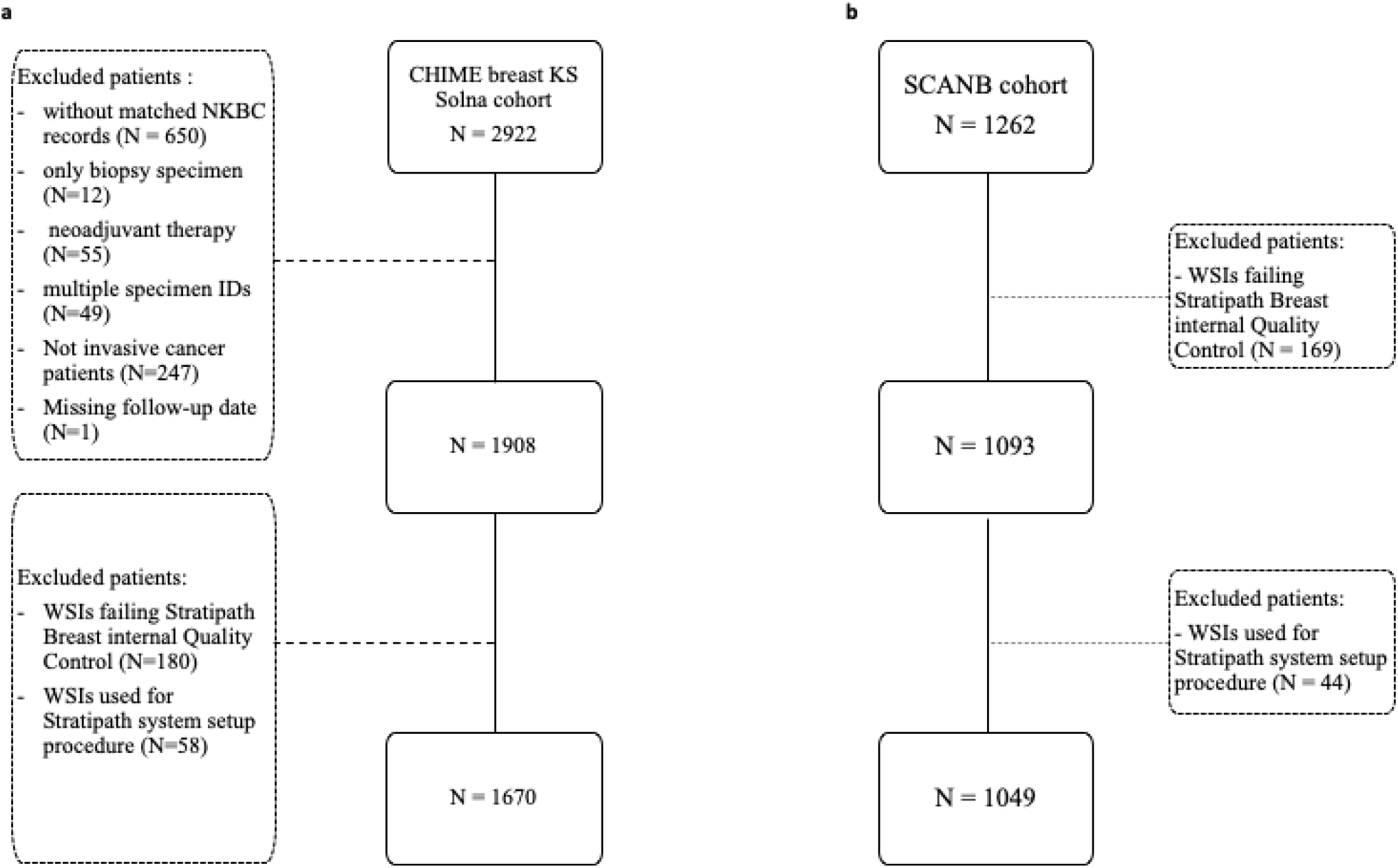
CONSORT diagram for the cohorts from two different sites in Sweden. **a**. CHIME breast KS Solna cohort with included patient whole slide images from Stockholm, Sweden. **b**. SCAN-B cohort with included patient whole slide images from Lund, Sweden. NKBC=National quality registry for breast cancer. WSI=whole slide image.

### Statistical analyses

We evaluated the prognostic performance of the patient risk groups (high risk and low risk) assigned by the Stratipath Breast (version 1.3) analysis. Progression-free survival (PFS) was the survival endpoint, defined as the time to local recurrence, distant metastasis or detection of contralateral tumours. The patients were followed from the initial date of diagnosis to the date of reported progression, or the last follow-up date (whichever came first). Kaplan-Meier (KM) curves were used to visualise the survival probability over time, for the risk groups assigned by Stratipath Breast, as well as for clinical NHG. The association between the different patient groups and PFS was assessed by estimating the Hazard ratio (HR) with 95% confidence interval (CI) using the Cox Proportional Hazard (PH) model. The multivariable Cox PH model was fitted by adjusting for established clinical covariates including: age at the time of diagnosis, tumour size, lymph node status, ER status, and HER2 status. Tumour size was converted to a binary covariate with tumour size <=20mm and >20mm. Lymph node status was defined as positive (metastasis to one or more lymph nodes) or negative (no lymph node metastasis). ER status was defined in accordance with local clinical guidelines, as the presence of >= 10% ER-positive cells in the immunohistochemistry (IHC) staining. HER2 status was defined according to national guidelines using IHC staining and for equivocal cases, gene amplification was confirmed by fluorescent or silver *in situ* hybridisation assays. Patients with missing clinical information were excluded from the analyses. Further, additional time-to-event analysis was performed by stratifying patients into five equally sized bins defined by quantiles in the continuous slide score from Stratipath Breast. The 5-quantile bins of the continuous slide score was created using the *quantcut* function in the package *gtools* (v.3.9.5) in R (20). All time-to-event analysis was performed using the *survival* package in R (21). P-value < 0.05 was considered as statistically significant in all the analyses. All statistical analyses were performed in R version 4.2.0.

## Results

### Prognostic patient stratification by tumour grade and Stratipath Breast

First, we assessed the prognostic stratification based on clinically assigned NHG, and the risk groups assigned by Stratipath Breast in all patients. We observed a significant difference in the PFS rate over time between the NHG groups (log-rank p-value < 0.0001) and between the Stratipath Breast low- and high-risk groups (log-rank p-value < 0.0001) (Figure 2a and 2b). We observed the adjusted HRs for PFS of 3.93 (95% CI: 1.41-10.99, p-value = 0.009) and 3.71 (95% CI: 1.26-10.90, p-value = 0.017) for NHG2 and NHG3 groups respectively with NHG1 as the reference (Figure 2c).The adjusted HR for PFS between the Stratipath Breast low- and high-risk groups was estimated to 2.64 (95% CI: 1.60-4.38, p-value < 0.001) (Figure 2d).

**Figure 2.**
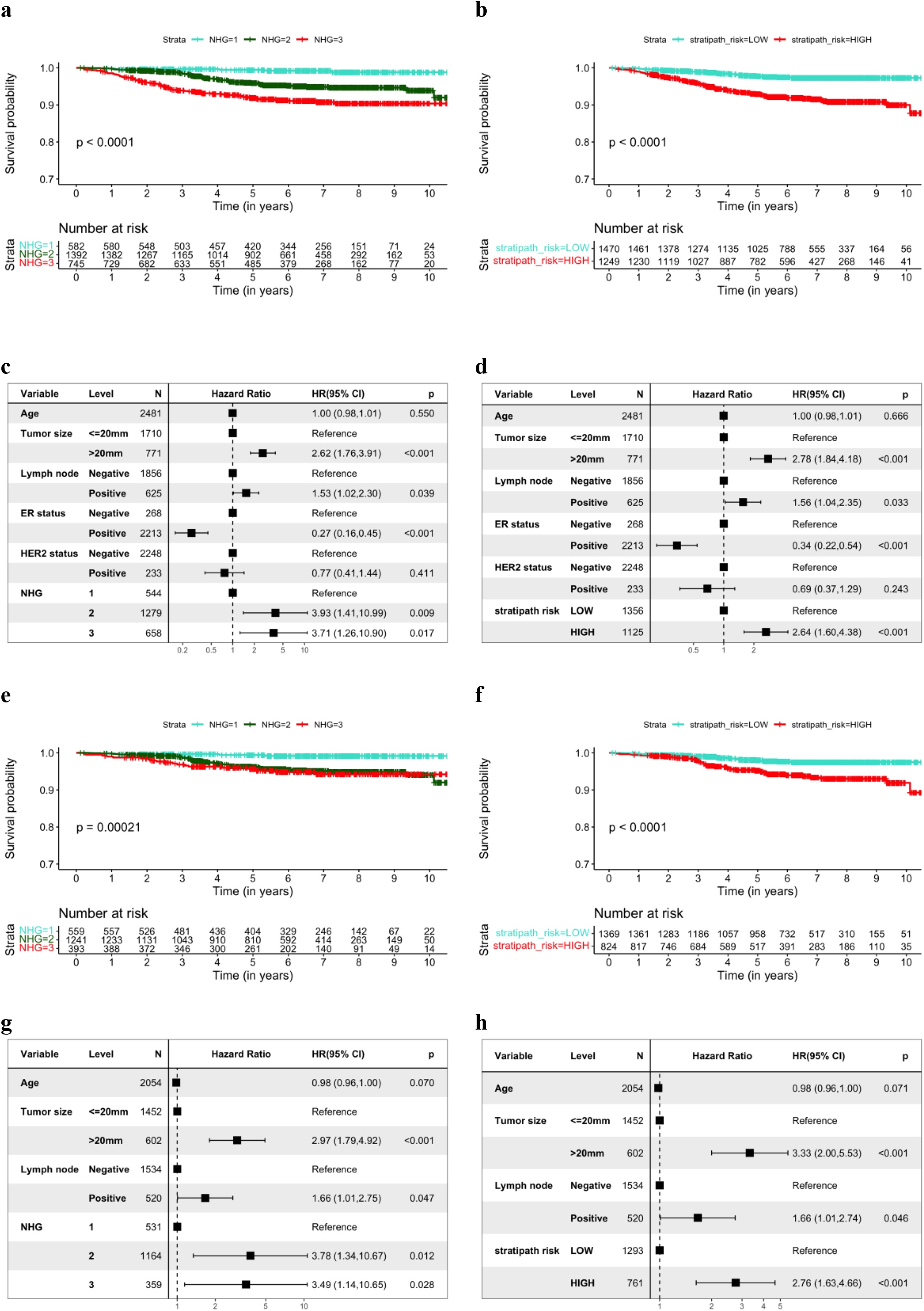
Prognostic stratification of patients by clinical NHG and Stratipath Breast. **a** Kaplan Meier (KM) curves for progression-free survival (PFS) among all patients stratified by clinical NHG. **b** KM curves for PFS among all patients stratified by Stratipath Breast risk groups. **c** Multivariable Cox Proportional Hazard (PH) regression estimating the association between NHG and PFS. **d** Multivariable Cox PH regression estimating the association between Stratipath Breast risk groups and PFS. **e** KM curves for PFS among ER+/HER2-patients stratified by clinical NHG. **f** KM curves for PFS among ER+/HER2-patients stratified by Stratipath Breast. **g** Multivariable Cox PH regression estimating the association between NHG and PFS in the ER+/HER2-patient subgroup. **h** Multivariable Cox PH regression estimating the association between Stratipath Breast risk groups and PFS in the ER+/HER2-subgroup. NHG=Nottingham histological grade, ER=oestrogen receptor, HER2=human epidermal growth factor receptor 2.

Next, we performed subgroup analysis in the ER+/HER2-patient subgroup, by clinical NHG and Stratipath Breast. We observed a significant HR for PFS between the NHG groups (log-rank p-value = 0.00021), as well as between the Stratipath Breast low- and high-risk groups (log-rank p-value < 0.0001) (Figure 2e and 2f). The adjusted HRs for PFS of 3.78 (95% CI: 1.34-10.67, p-value = 0.012) and 3.49 (95% CI: 1.14-10.65, p-value = 0.028) were observed for NHG2 and NHG3 groups respectively with NHG1 as the reference (Figure 2g). We observed an adjusted HR for PFS of 2.76 (95% CI: 1.63-4.66, p-value < 0.001) for Stratipath low-vs high-risk groups (Figure 2h).

### Prognostic performance in the subgroup of intermediate-risk patients

Next, we evaluated the prognostic performance of Stratipath Breast in the clinically relevant subgroup of intermediate-risk breast cancers. We investigated the NHG2 subgroup (Figure 3a), as well as the subgroup of NHG2/ER+/HER2-patients (Figure 3b) and observed significant difference in the PFS rate over time between the Stratipath Breast low- and high-risk groups in the KM curves (log-rank p-value < 0.0001) (Figure 3a and 3b). In multivariable Cox PH analysis, we observed a HR for PFS of 2.27 in the NHG2 subgroup (95% CI: 1.29-3.99, p-value = 0.004), and a HR of 2.20 (95% CI: 1.22-3.98, p-value = 0.009) in the NHG2/ER+/HER2-subgroup (Figure 3c and 3d).

**Figure 3.**
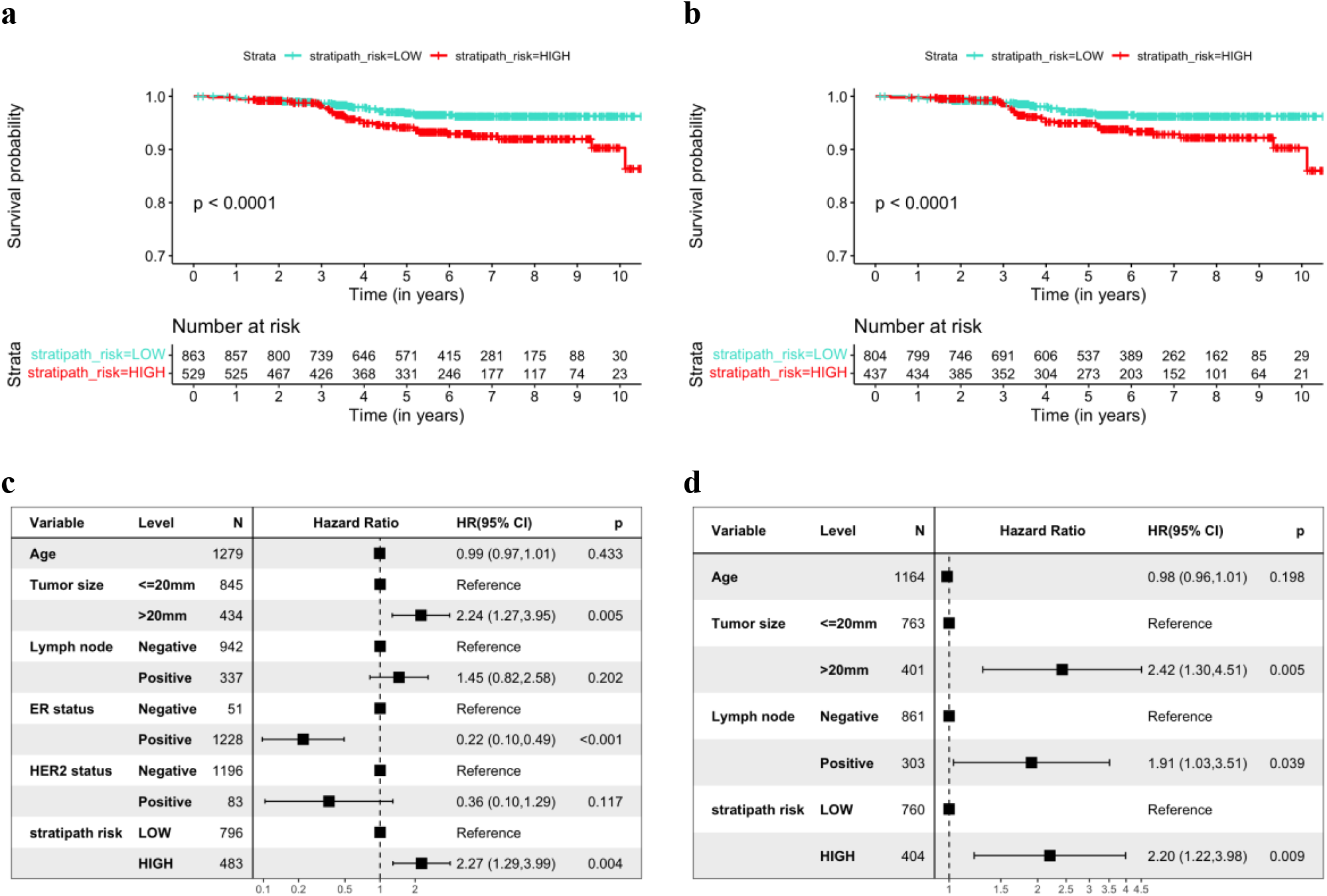
Prognostic stratification of patients with intermediate grade (NHG2) using Stratipath Breast with progression-free survival (PFS) as the outcome. **a** Kaplan Meier (KM) curves for PFS among NHG2 patients stratified by Stratipath Breast risk groups. **b** KM curves for PFS among NHG2/ER+/HER2-patients stratified by Stratipath Breast risk groups. **c** Multivariate Cox Proportional Hazard (PH) regression estimating the association between Stratipath Breast risk groups and PFS in NHG2 patients. **d** Multivariate Cox PH regression estimating the association between Stratipath Breast risk groups and PFS in the NHG2/ER+/HER2-patient subgroup. NHG=Nottingham histological grade, ER=oestrogen receptor, HER2=human epidermal growth factor receptor 2.

### Association of PFS with five level risk groups from Stratipath Breast

Lastly, we assessed the prognostic stratification of the Stratipath Breast continuous slide score by stratifying patients into risk groups based on five equally sized bins defined by quantiles in the slide score. We observed a significant difference in the PFS rate over time between the five level risk groups in all patients (log-rank p-value < 0.0001) and ER+/HER2-patient subgroup (log-rank p-value < 0.0001) (Figure 4a and Figure 4b). Further, for all patients, we observed an increase in HRs for PFS between the higher quantile risk groups and the lowest group (reference) in univariate Cox analysis (Figure 4c). In the ER+/HER2-patient subgroup, we did not observe the significant association of PFS with the second- and third-quantile risk group (p > 0.05), however, we did observe the significant association of PFS with the fourth- and fifth-quantile risk group with first-quantile risk group as the reference (p < 0.05) (Figure 4d). The adjusted HRs for fourth and fifth quantile were found to be similar and significantly associated with PFS in all patients and ER+/HER2-patient subgroup with first-quantile risk group as the reference (Figure 4e and 4f).

**Figure 4:**
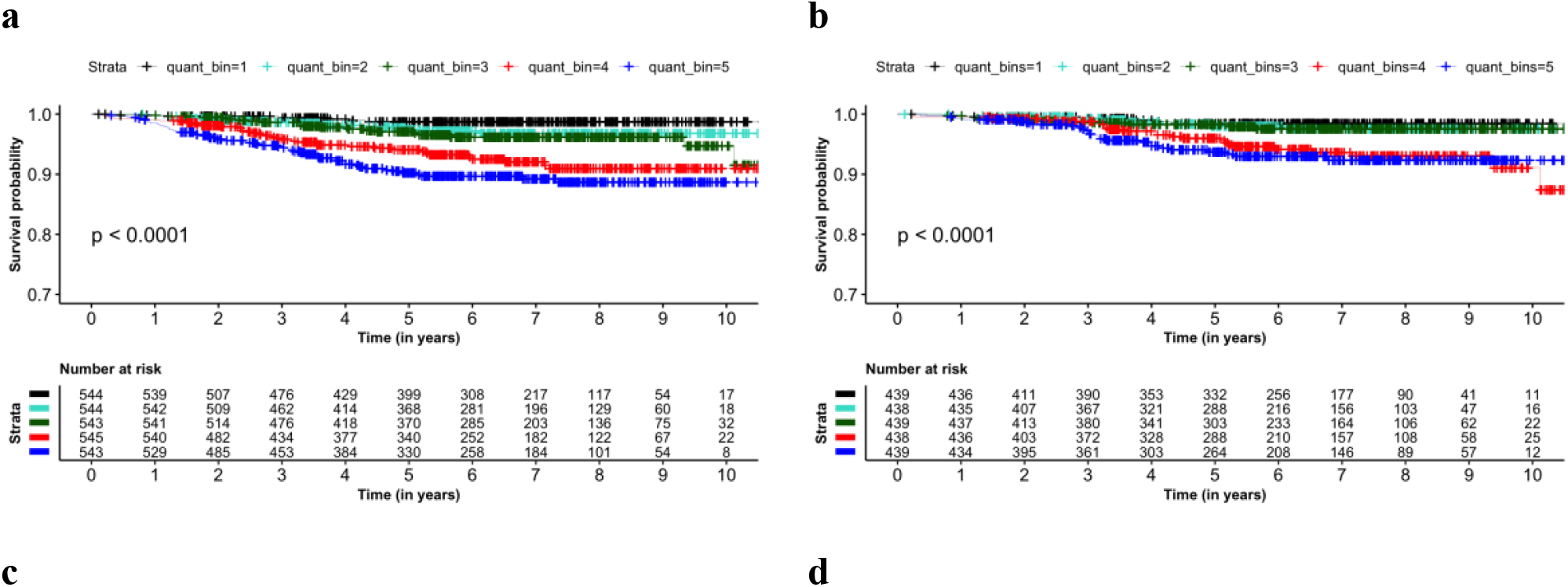
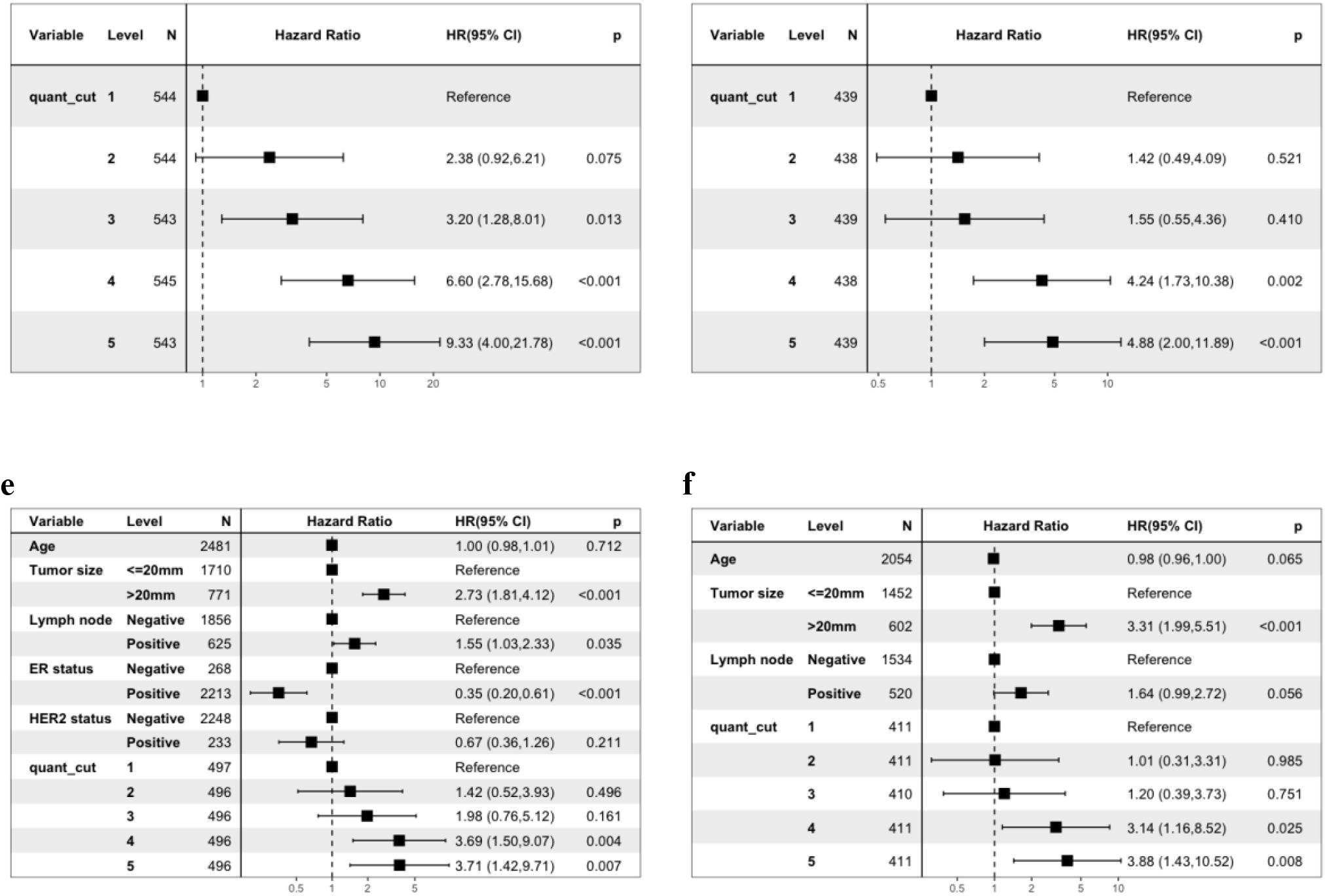
Prognostic stratification of the patients based on 5-quantile bins of the continuous slide score. **a, b** KM curves for progression-free survival (PFS) among all patients and ER+/HER2-subgroup stratified by 5-quantile groups respectively. **c, d** Univariate Cox PH regression estimating the unadjusted association of the five level risk groups and PFS for all patients and ER+/HER2-patient subgroup respectively. **e, f** Multivariate Cox PH regression estimating the association between the quantile risk groups and PFS in all patients and ER+/HER2-patient subgroup respectively. ER=oestrogen receptor, HER2=human epidermal growth factor receptor 2.

## Discussion

In this study, we validated the prognostic performance of the deep learning-based CE-IVD marked Stratipath Breast solution for risk stratification of invasive breast tumours from H&E-stained histopathology WSIs. We assessed the prognostic performance using PFS as the endpoint, in breast cancer patients from two different hospitals, for all patients as well as for the clinically relevant ER+/HER2-subgroup and the intermediate-risk subgroup.

Histological grading has a high inter-observer variability in clinical routine (3,22–24) and more than 50% of the patients in the ER+/HER2-patient subgroup are assigned as intermediate grade, NHG2 (3). However, the NHG2 assignment is not informative in clinical decision-making and is not directly related to the molecular characteristics of the tumours (25). Therefore, there is a need for assays or methods that can provide a refined risk stratification of the intermediate-grade patients to provide decision support when considering patients for adjuvant chemotherapy. Moreover, as a consequence of the low reproducibility in histological grading, it would be clinically valuable to enable more precise and reliable risk stratification of all breast cancer patients.

In terms of clinical value, the ER+/HER2-subgroup is not only the largest but also the group for which risk stratification is most valuable for therapy decision-making. In the other subgroups of breast cancer, namely HER2+ and triple negative, most patients are treated with adjuvant chemotherapy and/or targeted drugs. In the present study, we observed an adjusted HR for PFS of 2.76 (95% CI: 1.63-4.66) for Stratipath low-vs high-risk groups in the ER+/HER2-subgroup, and 2.20 (95% CI: 1.22-3.98) for the re-stratification of intermediate-grade (NHG2) patients in the ER+/HER2-subgroup. This is in line with Wang et al., which reported an adjusted HR for recurrence-free survival of 1.91 in the re-stratification of NHG2 patients into low- and high-risk groups among ER+/HER2-patients, using an academic model (research use only) in their independent external validation cohort (14). Furthermore, in this study we also evaluated prognostic performance in a five level risk group stratification based on the Stratipath Breast continuous score, and observed an adjusted HR for PFS of 3.88 (95% CI: 1.43-10.52) comparing the lowest risk group (reference) with the highest risk-group. The corresponding unadjusted HR for PFS was 4.88 (95% CI: 2.00-11.89). We note that groups 4 and 5 have approximately similar HR estimates in the ER+/HER2-subgroup, and also that the effect sizes observed in this study are similar to results reported for molecular-based assays based on gene expression profiling.

Several gene expression-based methods have been developed for clinical use to predict the risk of distant recurrence, especially for the selection of patients benefiting from adjuvant chemotherapy among ER+/HER2-node-negative early breast cancer patients. Popular methods include PAM50-based Prosigna ROR and Oncotype DX RS that stratify ER+/HER2-patients into low-, intermediate- and high-risk groups. Oncotype DX RS has been validated in retrospective analyses of prospective clinical trials (26,27) and prospectively in a prospective clinical trial (28–30). However, some of the retrospective comparative studies assessing the prognostic performance of the various multigene assays have reported better predictive performance of the overall and late distant recurrence using PAM50-based ROR score, Breast Cancer Index (11) and EndoPredict (EPclin) (31) over Oncotype DX RS in ER+/HER2-node-negative breast cancer patients (5,32). Most importantly, there is a poor correlation between the different gene expression profiling tests (33). Another limitation of the molecular profiling methods is the use of bulk tumour tissue analysis, in which intra-tumour heterogeneity is not accounted for (34). Furthermore, the effect of intra-tumour heterogeneity on the prognostic misclassification of the individual patients using multigene assays has been shown (35). In addition, both Prosigna and EPclin include clinical risk factors such as nodal status and tumour size in the risk profiling tests (31), which may impact the clinical interpretation, whereas Stratipath Breast does not incorporate any clinical parameters in the analysis. Instead those prognostic factors should be taken into account in the final comprehensive risk-assessment at the tumour board.

Three-risk group stratification from Oncotype DX RS showed an adjusted HR of 2.5 for distant recurrence (95% CI: 1.30-4.50) between intermediate-vs low-risk groups and a HR of 5.2 (95% CI: 2.7-10.1) for high-vs low-risk groups in the node-negative postmenopausal hormonal receptor-positive early breast cancer patients from the TransATAC study population (36). PAM50-based Prosigna ROR derived risk-groups reported adjusted HRs of 2.15 for distant recurrence (95% CI: 1.21-3.81, p=0.009) between intermediate-vs low-risk groups and of 4.26 (95% CI: 2.44-7.43, p<0.0001) for high-vs low-risk groups in the postmenopausal hormonal receptor-positive early breast cancer patients derived from the ABCSG-8 trial (4). Further, for the postmenopausal hormonal receptor-positive early breast cancer patients from the combined cohort derived from the combined TransATAC and ABCSG-8 trial, they observed HRs of 3.75 (95% CI: 2.19-6.41) for intermediate-vs low-risk and 5.49 (95% CI: 2.92-10.35) for high-vs low-risk groups in the HER2-node-negative patient subgroup with distant recurrence between 5 to 10 years as the survival endpoint (10). Importantly, the patients that were retrospectively derived from the clinical trials in the mentioned validation studies were randomised to the treatment arm (tamoxifen or anastrozole) and did not receive adjuvant chemotherapy. This could lead to a lower survival rate, especially among high-risk patients and might result in larger effect sizes for high-vs low-risk and intermediate-vs low-risk groups.

Furthermore, molecular profiling-based academic models have shown significant prognostic value in re-stratification of intermediate-grade patients into low- and high-risk groups (12,13). Sotiriou et al. developed the genomic grade index (GGI) based on the identified 97-gene signatures and observed an estimated HR of 3.61 associated with the risk of recurrence (12). Wang et al. reported an estimated HR of 2.43 associated with recurrence-free survival based on the 37 gene RNA-seq profiling for the re-stratification of intermediate-grade patients (13). Using AI-based image analysis, Stratipath Breast provides prognostic re-stratification based on digitised WSIs of H&E-stained breast tumours available in clinical routine without any further tissue preparation. Image-based risk stratification also provides the added benefit of short lead times and substantially lower cost compared to molecular diagnostics and therefore has the potential to reach broader patient populations.

Our study has several limitations. Since, the discussed multigene assays, except for EPclin, stratify patients into three-tier risk groups (low, intermediate and high) and have been validated in different cohorts, it is difficult to directly compare the prognostic performance with the binary stratification method (low and high) by Stratipath Breast. However, it remains an important question for the future to be answered, preferably with comparisons in the same study population. Another limitation of this study is the relatively short median follow-up time (6.25 years) in the present study.

In conclusion, in this study we demonstrate the prognostic performance of Stratipath Breast in independent data from two Swedish hospitals, using PFS as the endpoint. This AI-based image analysis tool provides a fast and cost-effective solution to provide precise prognostic patient stratification that can be used as a decision support tool in clinical decision-making, with the potential to contribute to the reduction of both over- and undertreatment of breast cancer patients.

## Data Availability

Data cannot be made public due to local legislation since it contains sensitive patient data.

## List of abbreviations

H&E: Haematoxylin & Eosin
WSI: Whole Slide Image
ER: Oestrogen Receptor
HER2: Human Epidermal Growth Factor Receptor 2
HR: Hazard Ratio
PFS: Progression-Free Survival
NHG: Nottingham Histological Grade
RS: Risk Score
ATAC: Arimidex, Tamoxifen Alone or in Combination
ROR: Risk Of Recurrence
ABCSG-8: Austrian Breast and Colorectal Study Group-8
GGI: Genomic Grade Index
AI: Artificial Intelligence
KM: Kaplan-Meier
IHC: Immunohistochemistry
CI: Confidence Interval
PH: Proportional Hazard

## Ethics approval and consent to participate

The study has approval from the Swedish Ethical Review Authority (2017/2106-31, with amendments 2018/1462-32 and 2019-02336).

## Consent for publication

Not Applicable.

## Availability of data and materials

Data cannot be made public due to local legislation since it contains sensitive patient data.

## Author’s contribution

AS performed statistical analysis and computing, visualisation, drafted and edited the manuscript. SKL, KLE and YW contributed to data analysis and editing of the draft manuscript. SR contributed to editing of the draft manuscript. JH conceived and conceptualised the study, contributed to preparing and editing of the draft manuscript, and provided resources and supervision. MR conceived, conceptualised and directed the study, contributed to preparing and editing of the draft manuscript, and provided resources and supervision. All authors approved of the final manuscript.

## Acknowledgements

The authors acknowledge patients, clinicians, and hospital staff participating in the SCAN-B study, the staff at the central SCAN-B laboratory at the Division of Oncology, Lund University, the Swedish National Breast Cancer Quality Registry (NKBC), Regional Cancer Center South and the South Swedish Breast Cancer Group (SSBCG). We also acknowledge the help and support from Dr Johan Vallon-Christersson at Lund University (Sweden) with the preparation of clinicopathological information for the SCAN-B study. SCAN-B was funded by the Swedish Cancer Society, the Mrs. Berta Kamprad Foundation, the Lund-Lausanne L2-Bridge/Biltema Foundation, the Mats Paulsson Foundation, and Swedish governmental funding (ALF). Stratipath AB provided the Stratipath Breast analysis.

