## Supplementary Table 1 for "Validation of an AI-based solution for breast cancer risk stratification using routine digital histopathology images"

**Table of content:**

Table 1. Baseline characteristics of the cohorts from two different hospitals in Sweden.

**Table 1:** Baseline characteristics of the cohorts from two different hospitals in Sweden.

| | CHIME breast KS Solna (N=1542) | SCAN-B (N=933) | Overall (N=2475) |
|---|---|---|---|
| **Age** | | | |
| Mean (SD) | 61.6 (12.3) | 65.5 (12.3) | 63.0 (12.5) |
| Median [Min, Max] | 63.0 [29.0, 94.0] | 70.0 [30.0, 95.0] | 65.0 [29.0, 95.0] |
| **Tumour size** | | | |
| <20mm | 1043 (67.6%) | 620 (66.5%) | 1663 (67.2%) |
| >=20mm | 486 (31.5%) | 302 (32.4%) | 788 (31.8%) |
| Missing | 13 (0.8%) | 11 (1.2%) | 24 (1.0%) |
| **Lymph node status** | | | |
| Negative | 1105 (71.7%) | 646 (69.2%) | 1751 (70.7%) |
| Positive | 311 (20.2%) | 260 (27.9%) | 571 (23.1%) |
| Missing | 126 (8.2%) | 27 (2.9%) | 153 (6.2%) |
| **NHG** | | | |
| 1 | 302 (19.6%) | 170 (18.2%) | 472 (19.1%) |
| 2 | 894 (58.0%) | 527 (56.5%) | 1421 (57.4%) |
| 3 | 346 (22.4%) | 236 (25.3%) | 582 (23.5%) |
| **ER status** | | | |
| Negative | 165 (10.7%) | 87 (9.3%) | 252 (10.2%) |
| Positive | 1370 (88.8%) | 842 (90.2%) | 2212 (89.4%) |
| Missing | 7 (0.5%) | 4 (0.4%) | 11 (0.4%) |
| **HER2 status** | | | |
| Negative | 1349 (87.5%) | 850 (91.1%) | 2199 (88.8%) |
| Positive | 151 (9.8%) | 70 (7.5%) | 221 (8.9%) |
| Missing | 42 (2.7%) | 13 (1.4%) | 55 (2.2%) |
| **PFS (in years)** | | | |
| Mean (SD) | 6.27 (2.53) | 5.24 (2.24) | 5.88 (2.47) |
| Median [Min, Max] | 6.49 [0.153, 10.8] | 5.23 [0.115, 10.2] | 6.00 [0.115, 10.8] |
| **PFS status** | | | |
| 0 | 1461 (94.7%) | 902 (96.7%) | 2363 (95.5%) |
| 1 | 81 (5.3%) | 31 (3.3%) | 112 (4.5%) |

NHG=Nottingham histological grade, ER=oestrogen receptor, HER2=human epidermal growth factor receptor 2, PFS=Progression-free survival.
